# Genetic estimates of correlation and causality between blood-based biomarkers and psychiatric disorders

**DOI:** 10.1101/2021.05.11.21257061

**Authors:** William R. Reay, Dylan J. Kiltschewskij, Michael P. Geaghan, Joshua R. Atkins, Vaughan J. Carr, Melissa J. Green, Murray J. Cairns

## Abstract

There is a long-standing interest in exploring the relationship between blood-based biomarkers of biological exposures and psychiatric disorders, despite their causal role being difficult to resolve in observational studies. In this study, we leverage genome-wide association study data for a large panel of heritable biochemical traits measured from serum to refine our understanding of causal effect in biochemical-psychiatric trait parings. In accordance with expectation we observed widespread evidence of positive and negative genetic correlation between psychiatric disorders and biochemical traits. We then implemented causal inference to distinguish causation from correlation and found strong evidence that C-reactive protein (CRP) exerts a causal effect on psychiatric disorders, along with other putatively causal relationships involving urate and glucose. Strikingly, these analyses suggested CRP has a protective effect on three disorders including anorexia nervosa, obsessive-compulsive disorder, and schizophrenia, whilst being a risk factor for major depressive disorder. Multivariable models that conditioned CRP effects on interleukin-6 signalling and body mass index suggested that CRP-schizophrenia relationship was not likely mediated by those factors. Collectively, these data suggest that there are shared pathways that influence both biochemical traits and psychiatric illness, including factors such as CRP that are likely to constitute a causal effect and could be targets for therapeutic intervention and precision medicine.

## INTRODUCTION

Psychiatric disorders arise from a complex interplay between genetic and environmental risk factors. Indeed, twin-based and genome-wide association (GWAS) studies’ heritability estimates have demonstrated the importance of genetic risk to the spectrum of psychiatric illness (1-3). In particular, GWAS have been successful in identifying regions of the genome associated with psychiatric disorders, as well as revealing both overlapping and distinct features amongst the genetic architecture of these traits (2-6). For example, our group previously demonstrated that several genes associated with schizophrenia were shared with other psychiatric disorders, along with genes that appeared more specifically linked to schizophrenia (4). The challenge for psychiatric genetics from here onwards is to integrate and expand these data such that the biological insights gained may be directly relevant for psychiatric practice.

GWAS has proven valuable beyond just gene discovery in psychiatry, in that it allows the study of relationships between sets of traits in terms of genetic correlation (7), as well as GWAS informed methods for causal inference (8,9). An area of continued interest is the interplay between circulating biochemical factors and the pathophysiology of psychiatric disorders (10–13). These studies have endeavoured to find biochemical traits readily detectable in blood, that, in theory, could be diagnostic or prognostic biomarkers for a given psychiatric disorder. Many of these hypotheses stem from the idea that peripheral biochemical traits may exert an effect on the brain, directly or indirectly through their effect on other mediators, and that the manifestation of mental illness is in part attributed to these factors which primarily act in the periphery (11,12,14–16). Identifying these biochemical-psychiatric relationships would be clinically valuable as many of these traits can be modulated by existing drugs and/or lifestyle interventions. However, progress in this field has been hampered by small sample size studies, along with the fact that the majority of these studies are observational in nature, and thus, likely subject to at least some confounding. Genetics offers an attractive prospect for studying biochemical traits in psychiatry as many such measures are heavily influenced by genetic factors, with germline genetic variants fixed at birth and immune to reverse causation in most instances. In this study, we attempt to harmonise inter-study variability by using a panel of large sample size (*N* > 300,000) biochemical GWAS from a single cohort (UK biobank) to investigate genetic overlap with different psychiatric disorders, along with putative causal effects. We found that the majority of biochemical traits tested were genetically correlated with at least one psychiatric trait, with evidence of convergent and divergent correlation profiles amongst the different disorders. Interestingly, we also demonstrated evidence that there may be a causal relationship on psychiatric phenotypes through circulating C-reactive protein (CRP), glucose, and urate – which may have direct implications for clinical practice.

## METHODS AND MATERIALS

### Psychiatric genome-wide association studies

GWAS summary statistics for nine European ancestry cohorts for the following disorders were obtained from the psychiatric genomics consortium (PGC): attention/deficit hyperactivity disorder (ADHD) (17), anorexia nervosa (AN) (18), autism spectrum disorder (ASD) (19), bipolar disorder (BIP) (20), major depressive disorder (MDD) (21), obsessive compulsive disorder (OCD), post-traumatic stress disorder (PTSD) (22), schizophrenia (SZ) (23), and Tourette’s syndrome (TS) (24). Given that cognitive symptoms are pervasively associated with psychiatric illness, we also included a GWAS of general cognitive ability (25). Further information regarding these studies is provided in the supplementary text.

### Blood-based biomarker genome-wide association studies

We obtained GWAS summary statistics for a series of blood-based biochemical traits from the large UK biobank (UKBB) sample performed by the Neale group (http://www.nealelab.is/uk-biobank). The key advantage of these data is its large sample size (N > 300,000) and that the biochemical traits analysed were obtained from a single large cohort. Specifically, we utilised a panel of 50 biochemical GWAS which had high or medium confidence estimates of SNP heritability that were significantly different from zero as outlined in supplementary table 1 and supplementary text. These traits included lipids, micronutrients, hormones, metabolites, and enzymes.

**Table 1:** The most significant genetic correlation between each trait and a biochemical GWAS.

| Biochemical trait | Psychiatric trait<sup>1</sup> | $r_g^2$ | SE | P |
| --- | --- | --- | --- | --- |
| High light scatter reticulocyte percentage | ADHD | 0.256 | 0.033 | $4.58 \times 10^{-15}$ |
| High light scatter reticulocyte count | OCD | -0.212 | 0.046 | $3.37 \times 10^{-6}$ |
| C-reactive protein | AN | -0.286 | 0.038 | $9.76 \times 10^{-14}$ |
| Vitamin D | ASD | -0.177 | 0.047 | $2 \times 10^{-4}$ |
| Creatinine | BIP | -0.106 | 0.027 | $7.23 \times 10^{-5}$ |
| Leukocyte count | Cognition | -0.198 | 0.024 | $2.66 \times 10^{-16}$ |
| Leukocyte count | MDD | 0.155 | 0.025 | $9.77 \times 10^{-10}$ |
| Leukocyte count | PTSD | 0.225 | 0.043 | $1.37 \times 10^{-7}$ |
| Lymphocyte count | SZ | 0.075 | 0.017 | $1.52 \times 10^{-5}$ |
| Lymphocyte count | TS | -0.093 | 0.039 | 0.017 |

<sup>1</sup>Psychiatric GWAS: attention/deficit hyperactivity disorder (ADHD), anorexia nervosa (AN), autism spectrum disorder (ASD), bipolar disorder (BIP), major depressive disorder (MDD), obsessive compulsive disorder (OCD), post-traumatic stress disorder (PTSD), schizophrenia (SZ), and Tourette's syndrome (TS).

### Genetic correlation

Genetic correlation between each psychiatric and biochemical trait was estimated using linkage disequilibrium score regression (LDSR) (7), with summary statistic cleaned (‘munged’) to contain around one million HapMap 3 SNPs outside the major histocompatibility complex (MHC) with minor allele frequency > 0.05 for consistency (https://github.com/bulik/ldsc). Briefly, LDSR estimates genetic covariance by regressing SNP-wise *χ*^2^, the product of the marginal SNP effects from both traits (*Z*_1_*Z*_2_), on its LD score, which is an estimate of total LD existing with that SNP. Trait hertiabilities are utilised to normalise the genetic covariance to obtain genetic correlation (*r_g_*). A key advantage of LDSR is that sample overlap only affects the LDSR intercept and not the slope, meaning we can accurately estimate *r_g_* between UKBB biochemical GWAS and psychiatric GWAS with UKBB samples included. We used the Bonferroni method to correct for the fifty traits tested. One biochemical trait was excluded from further analysis, Apolipoprotein B, as it exhibited negative heritability within the block jackknifing procedure to estimate the *r_g_* standard error in some instances. The resulting 49 x 10 matrix of LDSR *r_g_*, divided by its standard error to obtain *Z*, was subjected to a latent clustering method, finite Gaussian mixture modelling (GMM), with the mclust R package version 5.4.6 (26). We selected the most parsimonious clustering configuration based on parametrisation of the covariance matrix utilising the largest Bayesian Information Criterion (BIC) value.

### Biochemical polygenic scoring in a severe cognitive deficit subtype of schizophrenia

We sought to further investigate biochemical traits displaying psychiatric genetic correlation and examine their relevance to the clinical dimensions of psychiatric disorders. Specifically, we considered the heterogeneity of cognitive impairment observed in schizophrenia, wherein deficits often manifest before the first psychotic episode and are highly variable in their presentation throughout clinical course (27,28). We considered biochemical traits that were genetically correlated with either schizophrenia or cognition after the application of multiple testing correction and interrogated their relationship with severe cognitive deficit in a cohort of schizophrenia cases from the Australian Schizophrenia Research Bank (ASRB) cohort. The use of these data was approved by the University of Newcastle Human Research Ethics Committee (HREC) and the ASRB (28-31). Previously, Green *et al.* utilised multidimensional Grade of Membership (GoM) clustering with nine cognitive measures to derive subgroups of cognitive performance in the ASRB (28). The most parsimonious configuration were two clusters of schizophrenia cases termed cognitive deficit (CD), with more pervasive cognitive impairment, and cognitively spared (CS), displaying intermediate cognitive performance relative to CD cases and healthy controls. Polygenic scores (PGS) were constructed for the 25 biochemical traits correlated with either schizophrenia or cognitive ability in a genotyped subset of the ASRB with schizophrenia cases subtyped as CD or CS (N = 391, Supplementary materials). The full details of this cohort and the generation of PGS is described in the supplementary text. We tested the association of each biochemical PGS with CD status using binomial logistic regression covaried for sex and the first three SNP-derived principal components (Supplementary Materials). The variance explained (Nagelkerke’s *R^2^*) in the full model with the PGS versus the null (covariates and intercept only) model, was converted to the liability scale assuming a population prevalence for CD of 0.33% (32). The population prevalence for CD is somewhat arbitrary, however, given the population prevalence of schizophrenia is around 0.7%, and 43% of this portion of the schizophrenia cases in the ASRB cohort was subtyped as CD, we believe this was an appropriate value to select. A polygenic score for general cognitive ability and a schizophrenia polygenic risk score (PRS) was also derived in this cohort for comparison (Supplementary Materials).

### Latent causal variable models

Genetic correlation may reveal important insights into shared biology between two traits, however, this should not be interpreted as implying a causal relationship in either direction. To evaluate evidence for a causal relationship, we implemented the latent causal variable (LCV) model to estimate genetic causality between traits, as outlined extensively elsewhere (9,33). The LCV framework leverages the bivariate genome-wide distribution of marginal SNP effects on both traits to estimate *partial genetic causality*. Specifically, the LCV assumes a latent variable, *L*, mediates the genetic correlation between the traits, and tests the strength of the correlation of each trait with *L*. The mixed fourth moments (cokurtosis) of marginal effect sizes for each trait (*Z*_1_, *Z*_2_) are compared to evaluate the proportionality of effects on either trait. The main output of the LCV model is the posterior mean genetic causality proportion (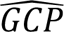), whereby 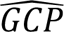 > 0 implies partial genetic causality of trait one on trait two, and *vice versa*. In other words, given 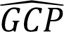 > 0, then trait one SNP effects 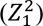 tend to be proportionally large on trait two (*Z*_1_*Z*_2_), such that 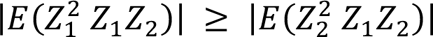. As in LDSR, the LDSR intercept is utilised here to guard against inflation due to sample overlap. We defined *partial genetic causality* using the recommended threshold of a significantly non-zero 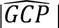 > 0.6, as this was shown by O’Connor and Price in simulations to guard against false positives (9). An LCV model was constructed for all genetically correlated psychiatric-biochemical trait pairs. Weak GCP estimates close to zero for genetically correlated traits imply that their relationship is potentially mediated by horizontal pleiotropy, whereby there are shared pathways, but the two traits do not likely exhibit vertical pleiotropy by acting within the same pathway. We also attempted to replicate the observed 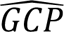 using a non-UKBB biochemical GWAS (34-37). It should be noted that the posterior mean 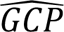 is not an estimate of the magnitude of any potential causal relationship and should not be interpreted as such – rather it evaluates the strength of evidence for a putative causal relationship using genome-wide SNP effects. The scripts to construct an LCV model are available from (https://github.com/lukejoconnor/LCV).

### Mendelian randomisation

C-reactive protein (CRP) exhibited strong evidence of partial genetic causality on multiple psychiatric disorders, and thus, we sought to estimate the magnitude of this causal relationship using univariable and multivariable Mendelian randomisation (MR). A detailed description of the MR methodology in this study is provided in the supplementary materials. MR is different from the LCV approach in that it leverages independent SNPs strongly associated with trait one (the exposure trait) as instrumental variables (IVs) to estimate the effect of the exposure on an outcome. The theoretical justification for using SNPs as IVs has been discussed extensively previously (8,38). We estimated the total effect of CRP on each disorder using independent genome-wide significant SNPs from a smaller non-UKBB GWAS as sample overlap between exposure and outcome can bias MR estimates (Supplementary Materials) (34). The *F*-statistic for IVs for this CRP GWAS was sufficiently strong (*F*-statistic > 10), as reported previously (34).

Our primary model was an inverse-variance weighted effect (IVW) estimator with multiplicative random effects, which assumes all IVs are valid and is less biased by heterogeneity than a fixed-effects IVW estimator (8). Whilst the IVW estimator is generally considered the most well-powered approach, the assumption that all IVs are valid is probably unrealistic in practice. As a result, we implemented a series of models which make different underlying assumptions regarding IV validity. Specifically, median based estimators which assume the majority of IVs are valid (39); a weighted mode estimator and a contamination mixture model, that both assume out of groups of IVs having the same asymptotic estimate, the largest group will be comprised of valid IVs (plurality valid) (40,41); and MR-Egger, which includes a non-zero intercept as a test of the average pleiotropic effect and assumes that there is no significant correlation between direct IV effects on the outcome and genetic association of IVs with the exposure (Instrument Strength Independent of Direct Effect (InSIDE) assumption) (42). As recommended by Bowden *et al.*, we ensured that the *I*^2^ statistic of IV-exposure effects exceeded 0.9, as this assesses the relative strength of the no-measurement error assumption, and thus, the suitability of using an MR-Egger model (43). We also tested the effect of using robust regression and penalised estimates for heterogeneity on the IVW, median, and Egger regression estimates (44). Evidence of horizontal pleiotropy and outliers were further investigated by testing heterogeneity in the IV/exposure-outcome effects (45), performing a leave-one-IV out analysis (38), testing for a non-zero MR Egger intercept (42), and an MR PRESSO test of global pleiotropy (also related to heterogeneity) (46). Furthermore, we also tested whether there was evidence of a causal effect in the reverse direction (psychiatric disorder as exposure), although MR estimates with binary exposures should be treated cautiously, as described elsewhere (47,48). Given only approximately 2,000,000 SNPs were available in the non-UKBB CRP GWAS utilised, we utilised the more deeply imputed UKBB CRP GWAS as the outcome here, although these results could therefore be inflated by sample overlap for AN and MDD.

Multivariable MR (MVMR) was then performed to evaluate the direct effect of CRP on each psychiatric outcome tested when conditioned on BMI and interleukin six (IL-6) signalling, which are both postulated to be closely functionally related to circulating CRP (Supplementary Text) (49,50). MVMR assumes that IVs are strongly associated with at least one exposure, and therefore, SNPs were chosen which were genome-wide significant for at least one phenotype. We constructed an IL-6 and BMI multivariable model separately – that is CRP conditioned on circulating IL-6 and its receptor (IL6R), and CRP conditioned on BMI, as well as BMI and IL- 6R. The strength of the IVs in each multivariable model was assessed using a two-sample conditional *F* statistic – which tests whether the IVs strongly predict each exposure, conditional on the other exposures in the model (51,52). When an *F* statistic > 10 could not be achieved, we relaxed the SNP inclusion threshold to represent suggestive significance in the GWAS (*P*_GWAS_ < 1 x 10^-5^, Supplementary Text). We compared direct estimates for each exposure using four different MVMR models – an IVW MVMR estimator, a median based MVMR estimator, an Egger regression based MVMR estimator, and a regularisation approach whereby LASSO type penalisation is applied to shrink intercept terms corresponding to IVs predicted as valid (MVMR-LASSO) (53). Given that CRP was genetically correlated with several psychiatric GWAS, genetic correlation may result in bias in MR estimates (9). However, MR is a valuable extension to the LCV model as it allows for specification of several different assumptions about IV validity and facilitates the estimate of total (univariable MR) and direct (MVMR) effects. The MR analyses were performed using the following packages in R version 3.6.0 – TwoSampleMR v 0.5.5 (54), MendelianRandomization v 0.5 (55), MRPRESSO v 1.0 (46), and MVMR v 0.3 (51).

### Genetic overlap between C-reactive protein and schizophrenia

We investigated whether any of the lead SNPs (genome-wide significant) reported in the PGC3 SZ GWAS were also associated with CRP (*P* < 5 x 10^-8^). For overlapping genome-wide significant signals, we tested whether there was a shared causal variant or different causal variants underlying these loci, assuming a single causal variant, using the *coloc* package colocalisation method (56). Moreover, we utilised *ρ*-HESS (https://github.com/huwenboshi/hess) to estimate local genetic covariance between SZ and CRP, as opposed to a genome-wide estimate by LDSR, with local genetic covariance (*ρ_g,local_*) calculated for munged HapMap3 SNPs in around 1,600 approximately independent LD blocks outside the MHC (57,58). Genes within the five LD blocks with the most statistically significant *ρ_g,local_* were subjected to pathway analysis using g:Profiler (https://biit.cs.ut.ee/gprofiler/gost) (59). We derived an estimate of genetic correlation (*r_g,local_*) by dividing *ρ_g,local_* by the product of the square roots of CRP and SZ local heritability per LD block, respectively.

### Downstream effects of C-reactive protein

We investigated the downstream effect of CRP on circulating levels of 3284 proteins in blood using MR (60). We used the larger UKBB CRP GWAS to select IVs to maximise power. The principal MR model was the IVW estimator with multiplicative random effects to maximise power. Proteins which demonstrated at least nominal association with CRP levels after multiple testing correction (FDR < 0.1) were investigated for protein-protein interaction and overrepresentation in biological pathways using STRING v 11.0 (61).

## RESULTS

### Widespread genetic correlation between blood-based biomarkers and psychiatric traits

We tested the genetic correlation between a panel of blood-based biomarkers and 10 psychiatric GWAS using LDSR. Interestingly, we found that 61% (N = 30) of the biochemical traits tested were significantly correlated with at least one psychiatric trait after multiple-testing correction, with every psychiatric trait exhibiting a significantly non-zero biochemical correlation after correction except for Tourette’s syndrome (Figure 1a, Supplementary Tables 2 – 11) The most significantly correlated biomarker for each trait is outlined in table 1.

**Figure 1.**
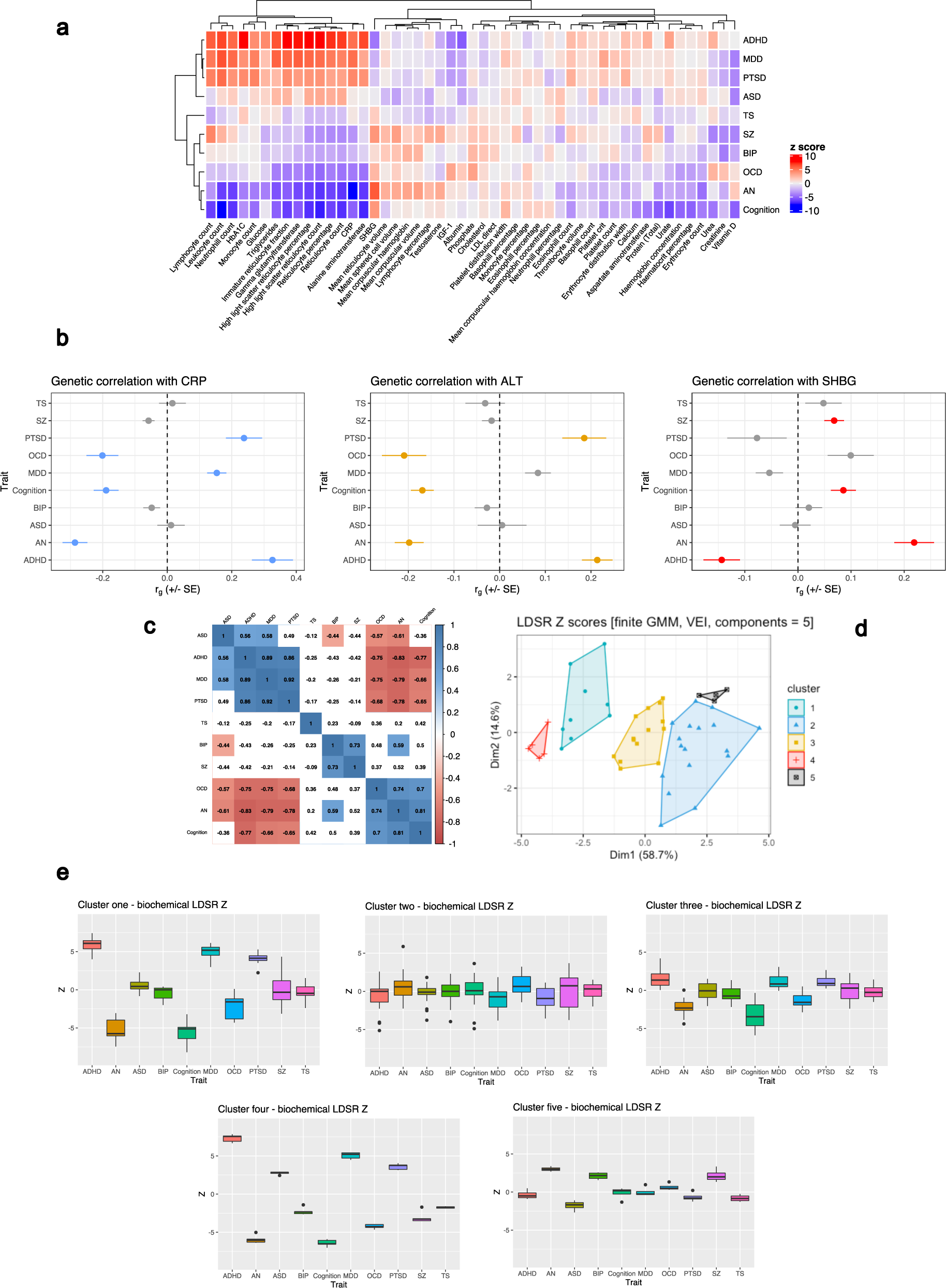
Genetic correlation between blood-based biomarkers and psychiatric GWAS. (**a**) Heatmap of LDSR correlation *Z* scores (*r_g_*/standard error) between each psychiatric trait and 49 biochemical GWAS. The psychiatric and biochemical traits are grouped on the *x* and *y* axes, respectively, by hierachial clustering using Pearson’s distance. (**b**) Examples of biochemical traits with evidence of discordant genetic correlations amongst the different psychiatric phenotypes. C-reactive protein (CRP), alanine aminotransferase (ALT), and sex-hormone binding globulin (SHBG) are presented for illustration. The forest plot denotes the LDSR *r_g_* with its standard error representing the confidence bars. Traits highlighted in blue, orange, and red, for CRP, ALT, and SHBG, respectively, were significantly correlated after the application of multiple testing correction. (**c**) Correlation matrix (Pearson) of LDSR *Z* between each trait, correlation estimates that survive correction for the number of tests performed are highlighted. (**d**) Components of biochemical LDSR *Z* scores derived using finite Gaussian mixture modelling (GMM) – the optimal parametrisation of the variance- covariance matrix was five components with diagonal distribution, variable volume, and equal shape (VEI). The components are plotted relative to their contribution to the first and second principal component of the LDSR *Z* matrix. (**e**) Box-and-whisker plots of the LDSR *Z* for each disorder composed of traits assigned to each of the five components derived from the GMM procedure.

**Table 2:** Strong evidence of partial genetic causality of a biochemical measure on a psychiatric trait.

| Biochemical trait | Psychiatric trait<sup>1</sup> | $GCP^2$ | $SE$ | $P^3$ | $r_g$ |
| --- | --- | --- | --- | --- | --- |
| Glucose | ADHD | 0.64 | 0.28 | 0.035 | 0.134 |
| C-reactive protein | AN | 0.92 | 0.07 | $5.95 \times 10^{-56}$ | -0.286 |
| C-reactive protein | MDD | 0.62 | 0.21 | $1.57 \times 10^{-12}$ | 0.154 |
| C-reactive protein | OCD | 0.75 | 0.16 | $2.07 \times 10^{-18}$ | -0.201 |
| Urate | Cognition | 0.88 | 0.09 | $5.27 \times 10^{-114}$ | -0.1047 |

<sup>1</sup>Psychiatric GWAS: attention/deficit hyperactivity disorder (ADHD), anorexia nervosa (AN), major depressive disorder (MDD), and obsessive-compulsive disorder (OCD).
<sup>2</sup>The posterior mean genetic causality proportion (GCP), with its accompanying posterior standard error in the adjacent column.
<sup>3</sup> $P$ value which tests whether the GCP estimate is significantly different from zero.

There was clear evidence of biomarkers with divergent genetic correlations between different psychiatric phenotypes – for instance, CRP, alanine aminotransferase (ALT), and sex-hormone binding globulin (SHBG, Figure 1b). In the case of CRP, it displayed a negative correlation after correction with OCD, AN, and cognition, as well as a trend towards a negative correlation with schizophrenia (*P* = 1.7 x 10^-3^), whilst its correlation with PTSD, MDD, and ADHD was strongly positive. These data provide some support to recent cohort studies, including lower CRP observed in patients with eating disorder (62), whilst elevated CRP was associated with ADHD (63). Notably, this contradicts previous observational estimates of elevated CRP in schizophrenia (64). Moreover, there were 14 biochemical traits which were only correlated after multiple testing correction with two or fewer psychiatric GWAS. A few such of examples of these more specific correlations included albumin and ADHD (*r_g_* = -0.151), mean corpuscular volume and AN (*r_g_* = 0.087), mean sphered cell volume and SZ (*r_g_* = 0.06), and creatinine with BIP and SZ (SZ: *r_g_* = -0.07, BIP: *r_g_* = -0.106).

We further investigated the relationship between the profile of 49 LDSR biochemical *Z* scores for each psychiatric trait. We observed strong positive and negative correlations between the trait-wise LDSR biochemical *Z* for each trait, which we term the *biochemical correlation profile* (Figure 1c). For instance, the ADHD biochemical correlation profile demonstrates large magnitude positive correlations with ASD, MDD, and PTSD but negative correlations with OCD, AN, and Cognition. This can be interpreted as traits which tend to be positively correlated with ADHD are also positively correlated with ASD, MDD, and PTSD and *vice versa* for OCD, AN, and Cognition. These relationships were further interrogated by subjecting the 10 biochemical correlation profiles to finite Gaussian mixture modelling (GMM, Figure 1d). We observed five components (clusters) with diagonal distribution, variable volume, and equal shape as the most parsimonious parameterisation of the covariance matrix (Supplementary Table 12). The biochemical correlation profile LDSR *Z* within each cluster are plotted in figure 1e. Briefly, the first component was composed of a series of traits with discordant correlations between the disorders, such as CRP, ALT, and glycaeted haemoglobin (HbA1c), whilst the second and third component were a diverse set of biomarkers with more similar LDSR *Z* across the psychiatric disorders tested. Component four was notable as it was solely composed of reticulocyte (immature erythrocytes) related traits, which analogous to component one was quite discordant in its correlation profiles. The fifth and final component was composed of other erythrocytic related traits, however, the differences between disorders were less marked than component four. Taken together, this demonstrates that groups of biomarkers tend to have similar relationships with different psychiatric traits.

### Genetically proxied biochemical measures were associated with a severe cognitive deficit schizophrenia subtype

We tested the association between 25 genetically proxied biomarkers (polygenic scores – PGS) that were correlated with either the schizophrenia or general cognitive ability GWAS and a severe cognitive deficit subtype of schizophrenia (Cognitive Deficit - CD) and a subset of cases with less marked impairment (Cognitively Spared – CS). There were two biochemical PGS that displayed a relatively significant association with CD status – haematocrit percentage and immature reticulocyte faction (Figure 2, Supplementary Table 13). Each standard deviation increase in the immature reticulocyte fraction PGS was associated with a 35.7% (95% CI: 14.5%, 56.9%, *P* = 4.72 x 10^-3^, *q* = 0.07) increase in the odds of severe cognitive deficit. Conversely, genetically proxied haemtocrit percentage exerted a protective effect – OR = 0.744 [95% CI: 0.533, 0.954], *P* = 5.82 x 10^-3^, *q* = 0.07. We emphasise that these signals only survive multiple testing correction using a lenient false-discovery rate (FDR) threshold of 10%, however, given the small sample size of this cohort, we believe that these findings remain noteworthy. Immature reticulocyte fraction was negatively correlated with cognition, with a trend towards a negative relationship with schizophrenia as well. Interestingly, haemtocrit percentage was also negatively correlated with cognition, however, there was a depletion of haemtocrit percentage alleles amongst CD vs CD schizophrenia cases, suggesting a more complex relationship may be present. Both the haemtocrit and reticulocyte fraction PGS explained around 1% of phenotypic variance in CD on the liability scale, which is similar to a PGS for general cognitive ability (Figure 2). A model constructed with all nominally CD-associated PGS (*P* < 0.05) explained almost 3% in phenotypic variance. There was a non-significant trend of enrichment of schizophrenia PRS (*P* = 0.054) in CD. Whilst these values are modest, it does suggest that the genetic architecture of biochemical traits is correlated with the severity of cognitive impairment in schizophrenia.

**Figure 2.**
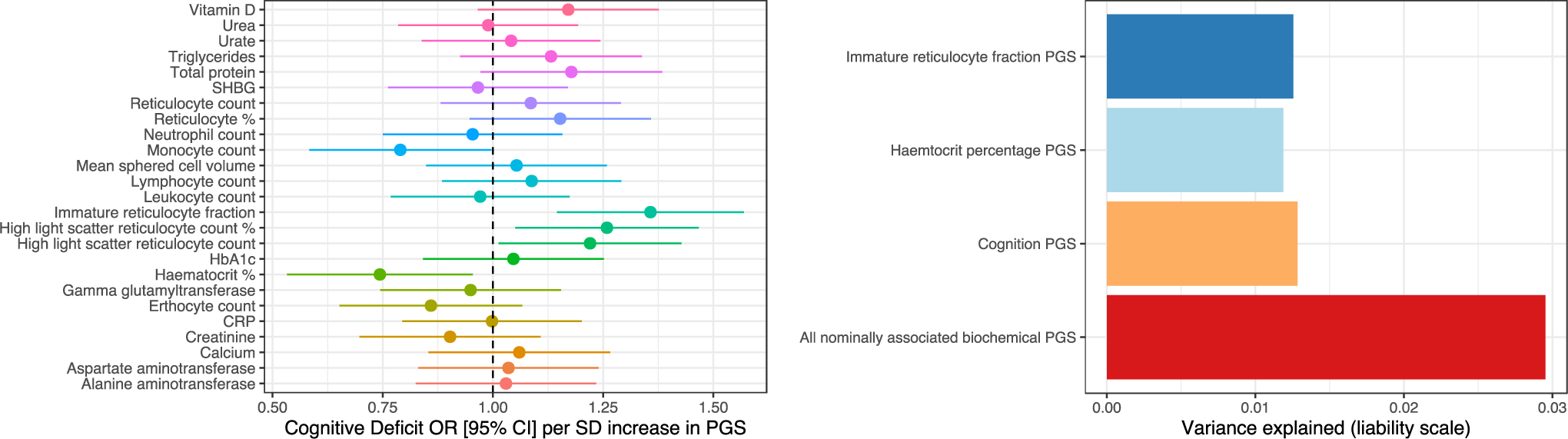
The association between biochemical polygenic scores and a severe cognitive deficit subtype of schizophrenia. Left panel is a forest plot of the association (odds ratio with 95% confidence interval) between a standard deviation increase in each polygenic score (PGS) a severe cognitive deficit (CD), relative to schizophrenia cases with less marked impairment (CS). The right panel is the variance explained on the liability scale (Nagelkerke’s *R*^2^, assuming an 0.33% CD population prevalence) for biochemical and a general cognitive ability PGS.

### Strong evidence of partial genetic causality between blood-based biomarkers and neuropsychiatric illness

A latent causal variable (LCV) model was constructed between each significantly correlated biochemical-psychiatric trait pair to estimate partial genetic causality (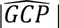 > 0.6 as strong evidence of partial genetic causality). We can then infer the consequence of the partial genetic causality of one trait on another using the sign of the genetic correlation. There were five instances where we found strong evidence of a potential causal relationship, with all of them suggesting an effect of the biochemical measure on the psychiatric trait, rather than *vice versa* (Table 2, Supplementary table 14). These were as follows: glucose on ADHD (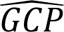 = 0.64), CRP on AN (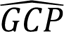 = 0.92), urate on cognition (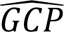 = 0.88), CRP on MDD (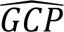 = 0.62), and CRP on OCD 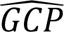 = 0.75). It is important to emphasise that these posterior mean (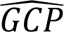 are not magnitudes of causal effect and only imply that there is a causal relationship between the biochemical traits and the psychiatric phenotype. Given the sign of the genetic correlation, we can likely infer that glucose may increase the risk of ADHD and urate could have a deleterious effect on general cognitive function. As outlined in a previous section, CRP has highly divergent correlations, and these data coupled with the LDSR further support a protective effect on AN and OCD, whilst it is likely risk increasing for MDD. SZ did not quite survive multiple- testing correction for a genetic correlation with CRP, however, given previous evidence of a protective effect of CRP on SZ from MR studies (65-67), we also constructed an LCV model between CRP and SZ and found moderate support for this relationship (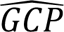 = 0.56, *SE* = 0.23, *P* = 5.11 x 10^-6^). Given the genetic correlation is only small it is less likely that previous MR studies were unduly biased by genetic correlation. In addition, we also observed an unusual phenomenon in the HbA1c and PTSD model, whereby the posterior mean (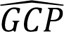 was strongly positive (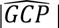 = 0.76), implying an effect of HbA1c on PTSD, whilst its *Z* score was negative (*Z* = -6.51), which signifies the opposite. As described in the supplementary material and supplementary figure 1, we found that these conflicting data was likely attributable to a rare violation of the LCV model assumptions, whereby the mixed fourth moments had opposite signs to each other and the genetic correlation. This could be explained by certain SNPs having highly divergent effects from the rest of the genome-wide signal. Moreover, there were two other trait pairs that trended towards partial genetic causation (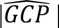 > 0.5) but did not exceed the stringent 0.6 threshold. These were SHBG on cognition (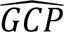 = 0.55) and triglycerides on OCD (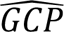 = 0.55). We then sought to replicate the LCV findings by utilising different previously published GWAS for glucose, urate, CRP, and HbA1c. Despite smaller sample sizes, we found relatively consistent GCP estimates which supported the above models using the UKBB GWAS (supplementary table 14). The exception to this was HbA1c, which did not show strong evidence of partial genetic causality (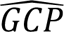 = 0.13) using a smaller sample size GWAS by Wheeler *et al.* (37).

### C-reactive protein levels exert a direct protective effect on schizophrenia conditioned on body mass index and interleukin-6 signalling

CRP displayed strong evidence of partial genetic causality on three psychiatric disorders, and thus, we sought to further analyse these relationships by estimating total effects and direct impact of CRP using univariable and multivariable Mendelian randomisation (MR), respectively (Supplementary Tables 16-22). We included SZ in these analyses, due to evidence from previous SZ GWAS that CRP exerts a protective effect on schizophrenia liability, as discussed in the previous section. The CRP GWAS utilised here was drawn from a non-UKBB cohort such that there was no sample overlap with the AN and MDD GWAS (Supplementary Materials) (34). Using our primary model (IVW estimator with multiplicative random effects), we found evidence that a natural log transformed mg/L increase in CRP was associated with a statistically significant reduction in the odds of SZ (OR = 0.91 [95% CI: 0.85,0.98], *P* = 0.01) and AN (OR = 0.91 [95% CI: 0.83,0.99], *P* = 0.03), which supports the LDSR inferred direction of the LCV relationship. Using a less conservative IVW estimator with fixed effects yielded a more precise estimate in both instances, as expected (CRP → SZ *P* = 6.29 x 10^-5^, CRP → AN *P* = 9.79 x 10^-3^). There was a trend towards an odds increasing effect of CRP on MDD (*P* = 0.19), whilst there was no indication of a reliable effect in the OCD model (*P* = 0.82). It should be noted that as MR exploits independent (relative linkage equilibrium) genome-wide significant variants as IVs, as opposed to using genome-wide marginal effects in the LCV approach, and thus, a non-significant estimate from MR does not necessarily preclude the existence of a causal relationship, although LCV models with corresponding MR support would perhaps be viewed as stronger evidence. We discuss the sensitivity analyses for each of these univariable models in detail in the supplementary text. Briefly, the five MR tests deployed with different assumptions regarding IV validity (plurality valid, majority valid, and InSIDE assumption) had very similar point estimates (OR range: 0.88-0.91) and were all statistically significant for CRP → SZ with the exception of the simple median (*P* = 0.07). The CRP → AN estimate across the different models were also directionally consistent, however, they were not statistically significant (except for some contamination mixture models with different prespecified standard deviation of invalid IVs), and thus, the total effect of CRP on AN by MR has comparatively weaker evidence compared to SZ. The effect of using robust, penalised, or robust penalised weights in the median, IVW, and Egger models was not marked for each of the CRP psychiatric models (Figure 3a, Supplementary Table 21). In the CRP → SZ and CRP → AN model, the Egger intercept was not significantly different than zero (although there was a trend in the AN model, *P* = 0.07), and thus, there was no strong statistical evidence of confounding pleiotropy using this metric. However, there was evidence of heterogeneity between the IV ratio estimates for AN and SZ: the MR PRESSO global test of pleiotropy was significant and Cochran’s *Q* statistic significant for AN and SZ, although both causal estimates remained statistically significant in the MR-PRESSO outlier corrected estimates (Supplementary table 19). Given the biological complexity of these phenotypes, heterogeneity does not necessarily imply confounding pleiotropy. Moreover, using a leave-one-out-analysis, we found evidence of one outlier IV in the SZ model, whilst there were three outlier SNPs in the AN model (Supplementary Table 20), although the effects of removing these IVs were relatively small. The outlier IV in the SZ model was also proximal to the CRP gene itself (IVW *P* = 0.07 when removed), meaning it is likely to influence SZ through CRP, rather than being indicative of confounding pleiotropy. Given the CRP estimates on AN and SZ remained significant removing outliers by MR PRESSO or through iterative single IV exclusion, it is less likely that horizontal pleiotropy fully explains these signals. There was also no evidence using a reverse MR model, with the psychiatric disorder as the exposure, that there were bidirectional effects, although these models are inherently underpowered and are best treated as a test of the null hypothesis only (Supplementary Table 22).

**Figure 3.**
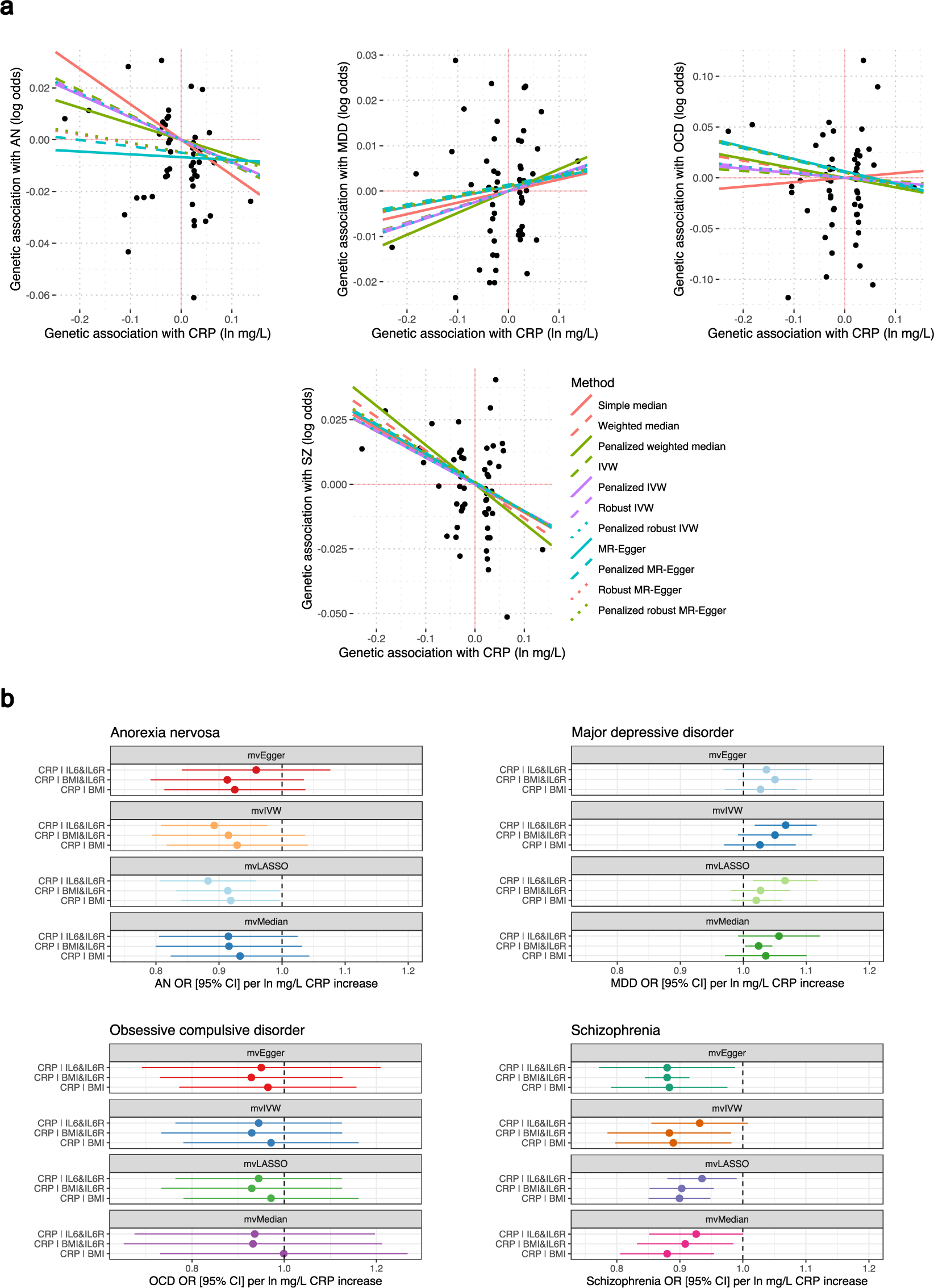
Total and direct estimated effect of C-reactive protein on psychiatric illness. (**a**) Total effect of CRP on each psychiatric outcome considered. Each point represents the IV-exposure vs IV-outcome effect. Trend lines are indicative of the slope of each MR method utilised. The outcomes plotted from left to right are AN, MDD, OCD, and SZ. (**b**) Direct effect of CRP in multivariable MR models – the three models contained the following phenotypes as additional exposures: i) circulating interleukin 6 (IL6) and its receptor (IL6R), body mass index (BMI), and IL6R and BMI.

We sought to investigate the direct effect of circulating CRP on the above psychiatric phenotypes using multivariable Mendelian randomisation (MVMR) conditioning on IL-6 signalling and BMI. We found that CRP exhibited a robust direct protective effect on schizophrenia conditioned on IL-6 signalling and BMI (Supplementary table 23, Figure 3b). For instance, using a multivariable IVW estimator, the direct effect of CRP on schizophrenia conditioned on BMI and IL-6R was analogous to the univariable IVW total estimate (OR = 0.88 [95% CI: 0.79, 0.98], *P* = 0.01). There was no evidence of an effect of BMI or IL-6 signalling on schizophrenia conditioned on CRP. Similarly, the effect size of the CRP → AN MVMR models remained similar to that estimated in the univariable constructs (Figure 3b), however, these estimates were only statistically significant in a subset of the models (Supplementary table 23). As a result, the MR evidence for the direct protective effect of CRP on AN is comparatively weaker than the CRP → SZ model, as was seen in the univariable estimates. We also observed some evidence to suggest that IL-6 abundance exerts a protective effect on AN conditional on CRP and IL-6R, which was nominally non-zero in every MVMR model except for the median estimator (OR = 0.96 [95% CI: 0.93, 0.99] per unit increase in IL-6, *P* = 0.037 – multivariable IVW-multiplicative random effects). Interestingly, whilst evidence for a direct effect of CRP on MDD conditioned on each variable set was weak (Figure 3b, Supplementary Table 23), there was consistent evidence that elevated IL-6R was associated with increased odds of MDD conditioned on CRP and IL-6 or CRP and BMI. For example, each unit increase in blood IL-6R protein expression was estimated to increase the odds of MDD by 2.7% [95% CI: 0.6%, 4.9%] conditioned on CRP and BMI. Finally, there was no evidence in the MVMR models to support the protective effect of CRP on OCD, however, there was evidence that increased BMI decreases the odds of OCD conditioned on CRP and IL-6 signalling (Supplementary Table 23). This BMI → OCD effect estimate was quite large, albeit with wide confidence intervals – OCD OR per SD increase in BMI conditioned on CRP and IL-6R = 0.52 [95% CI: 0.14, 0.91], *P* = 1 x 10^-3^.

### C-reactive protein displays overlapping association signals with schizophrenia and may have downstream impacts on the brain

There were five schizophrenia GWAS lead SNPs (*P* < 5 x 10^-8^), that also obtained genome-wide significance in the UKBB CRP GWAS. Colocalisation analyses using the European only subset of the SZ GWAS demonstrated strong evidence for the association of three of these loci with both SZ and CRP, however, there was likely a different underlying causal variant (Supplementary Table 24, Supplementary Text, Supplementary Figure 2). Local genetic correlation estimates with *ρ*-HESS demonstrated that 13 of the LD block partitions of the human genome displayed non-zero local covariance between CRP and SZ (Supplementary Table 25, Supplementary Figure 3) after Bonferroni correction. Interestingly, five of these LD blocks with strong evidence of local genetic covariance were positive, and thus, for these regions of the genome SZ and CRP were positively correlated in contrast to the genome-wide estimate of nominal negative correlation. We investigated the pathways overrepresented for genes physically mapped to the LD blocks of the top five most significant *r_g,local_* estimates (Supplementary Table 27). Interestingly, we found that *retinol metabolism* was the most significantly overrepresented gene-set after multiple testing correction, which is notable given evidence that retinoids play a role in the pathogenesis of SZ (30,68).

We also sought to investigate the downstream consequences of raised CRP. We estimated the effect of raised CRP on the expression of 3284 proteins in blood using univariable MR. Using a liberal FDR cut-off of 10%, we found that 95 proteins were putatively causally influenced by elevated CRP, with 45 of these proteins surviving a stricter FDR threshold of 5% (Supplementary Table 28). We emphasise that these analyses are exploratory in nature and we treat the effect sizes of the CRP effect on each protein largely as a test of the null hypothesis that the two are not associated. The vast majority of proteins prioritised using the IVW estimator were directionally consistent in the median, mode, and Egger sensitivity analyses (Supplementary Table 29). However, 20 of the FDR < 0.1 proteins were suggested to act in the opposite direction and causally influence CRP as the estimated variance explained by the IVs was significantly larger in the outcome than the CRP exposure, although this does not rule out bidirectional effects (Supplementary Table 30). We found that these proteins putatively influenced by CRP were enriched in several pathways including glycaemic signalling and lymphocyte biology (Supplementary Tables 31-33). Notably, there were neuronal pathways overrepresented with CRP-associated proteins – including, *axon guidance*, *dopaminergic synapse*, *neurogenesis*, *glial cell differentiation*, and *cholinergic synapse*. For example, dopaminergic and cholinergic synapse overrepresentations were driven by the three genes (*AKT1*, *AKT2*, and *AKT3*) that encode the RAC-alpha/beta/gamma serine/threonine-protein kinase complex, which was measured as a single entity in the protein study. Collectively, these data provide preliminary evidence that CRP may influence the expression of neuronally relevant proteins, and subsequent studies should seek to investigate these associations and their significance for different psychiatric phenotypes.

## DISCUSSION

In this study, we investigated genetic correlation and causality between a diverse panel of biochemical traits and psychiatric disorders, as well as general cognitive ability and a cognitive deficit subtype of schizophrenia. Our data demonstrated that there is clear evidence of genetic overlap between blood-based measures and psychiatric phenotypes, as quantified by LDSR, which is likely indicative of shared variants and pathways that predispose to these traits. Interestingly, the distribution of biochemical-psychiatric correlations demonstrated traits often exhibited highly divergent correlations (different signs), as well as clusters of biochemical measures that tended to have similar psychiatric correlation profiles. For instance, we found that five reticulocyte traits clustered together (Figure 1e), and they tended to have opposing psychiatric correlations (strong positive correlation with ADHD, MDD, and PTSD, whilst negative correlations with AN, OCD, SZ, and cognition). The genetic architecture of reticulocyte related traits remains relatively uncharacterised, however, it is a highly polygenic trait that also has demonstrated genome-wide significant associations with rare non-synonymous variation in genes such as *SPTA1*, *E2F4*, and *IFRD2* (69,70). Future study should further investigate how the genetic factors which contribute to reticulocyte biology may also influence psychiatric traits. Given the LCV posterior mean GCP estimates between the genetically correlated reticulocyte traits and each psychiatric phenotype were low, it suggests the existence of horizontal rather than vertical pleiotropy.

A key advantage of our study is that we extended the findings from the LDSR models to estimate which biochemical-psychiatric trait pairs may represent causal relationships. We caution that all of these findings require validation in well-powered, replicated, randomised controlled trials to confirm that the causal effects do indeed exist. The putative effect of urate and glucose on cognition and ADHD, respectively, may have direct implications for drug repurposing given that compounds which modulate these traits are readily available. Both of these relationships are also supported by previous observational data (71-73). Indeed, data from an incident cohort study which treated individuals with urate lowering compounds allopurinol and febuxostat demonstrated evidence of a risk decreasing effect on dementia, supporting the deleterious effect of urate on general cognitive function (74).

The inferred effect of CRP on different psychiatric disorders was particularly interesting given that there was evidence of an odds-decreasing effect on AN, OCD, and SZ, whilst the opposite is true for MDD. CRP is traditionally conceptualised as a biomarker of chronic inflammation; however, its biology is likely somewhat more complex given it is also implicated to play a direct role in pathogen response (75,76). Moreover, as reviewed by Del Giudice and Gangestad, CRP in its hepatically secreted pentameric isoform demonstrates some anti-inflammatory effects and may be a marker of other non-inflammatory states (77). In our study, we also provide evidence that CRP levels may also exert an effect on proteins with neurological significance, including proteins enriched in pathways relevant to psychiatric illness such as axon guidance. Moreover, we show for the first time the previously documented protective effect of CRP on SZ through MR is not likely attributable to the effect of BMI and IL-6 signalling, which are closely related variables to CRP. Data from our study and previous examinations of the relationship between CRP and SZ through MR seemingly contradict previous observational evidence that CRP is elevated in SZ (64,78). If we assume that there is a causal effect, there are a number of explanations that could account for this, although all require further investigation. Firstly, previous observational studies that directly measured CRP in case/control cohorts could be confounded due to a variety of variables including lifestyle and general health, or even be caused by factors related to psychosis and/or schizophrenia itself. Whilst there was no evidence in this study using the LCV model or reverse MR that SZ causally influences CRP levels, local significant estimates of positive and negative genetic correlation between SZ and CRP observed for several LD blocks suggests a complex interrelationship between these phenotypes may exist that warrants further study. Secondly, CRP itself may influence factors that impact the brain, directly or indirectly, and these factors may play a role in the pathogenesis of psychiatric illness. For instance, CRP was postulated in this study to causally upregulate the expression of the RAC-alpha/beta/gamma serine/threonine-protein kinase complex (*AKT1*, *AKT2*, and *AKT3*), with impaired signalling by these serine/threonine kinases implicated to dysregulate dopaminergic neurotransmission and downregulation of genetically predicted neuronal *AKT3* expression associated with SZ via a transcriptome-wide association study (23,79). Finally, given that infection has been associated with liability of SZ, the role of CRP in pathogen defence may contribute to its putative protective properties. Further study on the neurobiological consequences of CRP signalling and its role in SZ is warranted, particularly to reconcile the discrepancies between observational studies and MR.

The putative causal effect of CRP on AN was demonstrated in this study is, to our knowledge, a novel finding; however, it does support data from a recent longitudinal study which demonstrated that elevated CRP was associated with a protective effect on eating disorders (62), along with decreased measured CRP observed specifically in AN (80). It should be noted that whilst the LCV data supported strong partial genetic causality of CRP on AN, the MR evidence was less statistically significant, with some evidence in the MVMR that IL-6 signalling may exert a protective effect on AN conditioned-on CRP. Inhibition of the IL-6 pathway has been associated with weight gain which may be protective for AN (81). Furthermore, CRP likely is intertwined with other metabolic factors, including insulin signalling, which our group has previously shown through MR is also putatively protective for AN (48). In MDD, we found some evidence to support that upregulation of the IL-6 receptor may be risk increasing conditioned on CRP and BMI, supporting preliminary data that blockade of IL-6R by agents like tocilizumab may improve depressive symptoms (82). However, data related to the anti-depressant qualities of tocilizumab are conflicting (83), and randomised control trials are warranted to further investigate repurposing opportunities for IL- 6 inhibition in MDD. Finally, BMI demonstrated a quite robust protective effect on OCD conditioned on CRP and IL-6 signalling, in accordance with observational data that OCD is associated with reduced odds of obesity (84).

There are a number of important limitations that are central to the interpretation of the data in this study. Genetic correlations from LDSR likely reflect a shared underlying genetic architecture, however, this could be mediated by the relationship of the same genetic variants to another variable or variables (7). Despite this limitation, the existence of horizontal pleiotropy between traits is still informative as identifying genes which effect both psychiatric and biochemical traits, and further insight into the mechanisms, would likely refine our understanding of both traits. Moreover, the genetically informed causal inference approaches we implement in this study are subject to limitations regarding the data they are performed with and any biases therein, including potential effects of population stratification (85), selection bias (86), and the assumption of acyclicity. The LCV model is also fixed to be bivariate in nature, and thus, the effects of multiple meditators cannot be taken into account. We address this by constructing multivariable MR models such that direct effects are estimated conditioned on likely confounders, however, our selection of confounders is not exhaustive, and other unidentified factors may influence our findings. The UKBB sample is also composed of older individuals over the age of 40, and thus, more developmentally sensitive effects on the biochemical traits in question could not be assessed. Genetic variants are also sometimes claimed to represent a lifetime effect on a particular variable, although caution is required in this inference given that factors like age may modulate the effect of a variant (87,88). We assert that although GWAS informed causal inference has a number of caveats and limitations it enables an important opportunity to priortise biochemical traits which are putatively clinically relevant in psychiatry and inform future study into these traits.

## Data Availability

All GWAS summary statistics are freely available without restriction, with the source of each GWAS fully described in the manuscript. ASRB data are available upon application (https://www.neura.edu.au/discoveryportal/asrb/).

## ACKNOWLEDGEMENTS

The authors wish to acknowledge Dr Luke O’Connor for his insightful correspondence in relation to some of the LCV data in this study. This work was supported by a National Health and Medical Research Council (NHMRC) grant (1188493). M.J.C. is supported by an NHMRC Senior Research Fellowship (1121474), URL: https://www.nhmrc.gov.au/. The funders had no role in study design, data collection and analysis, decision to publish, or preparation of the paper.

## DISCLOSURES

The authors declare no competing financial interests

## AUTHOR CONTRIBUTIONS

W.R.R. conceived and designed the study with input from M.J.C. W.R.R. performed the primary analyses. M.P.G and D.J.K performed the rho-HESS analyses. J.R.A performed the SNP imputation in the ASRB cohort. M.J.C, V.J.C, and M.J.G contributed the ASRB cohort, as well as to critical interpretation of the results and drafting of the manuscript.

---

**Supplementary Figure 1.**
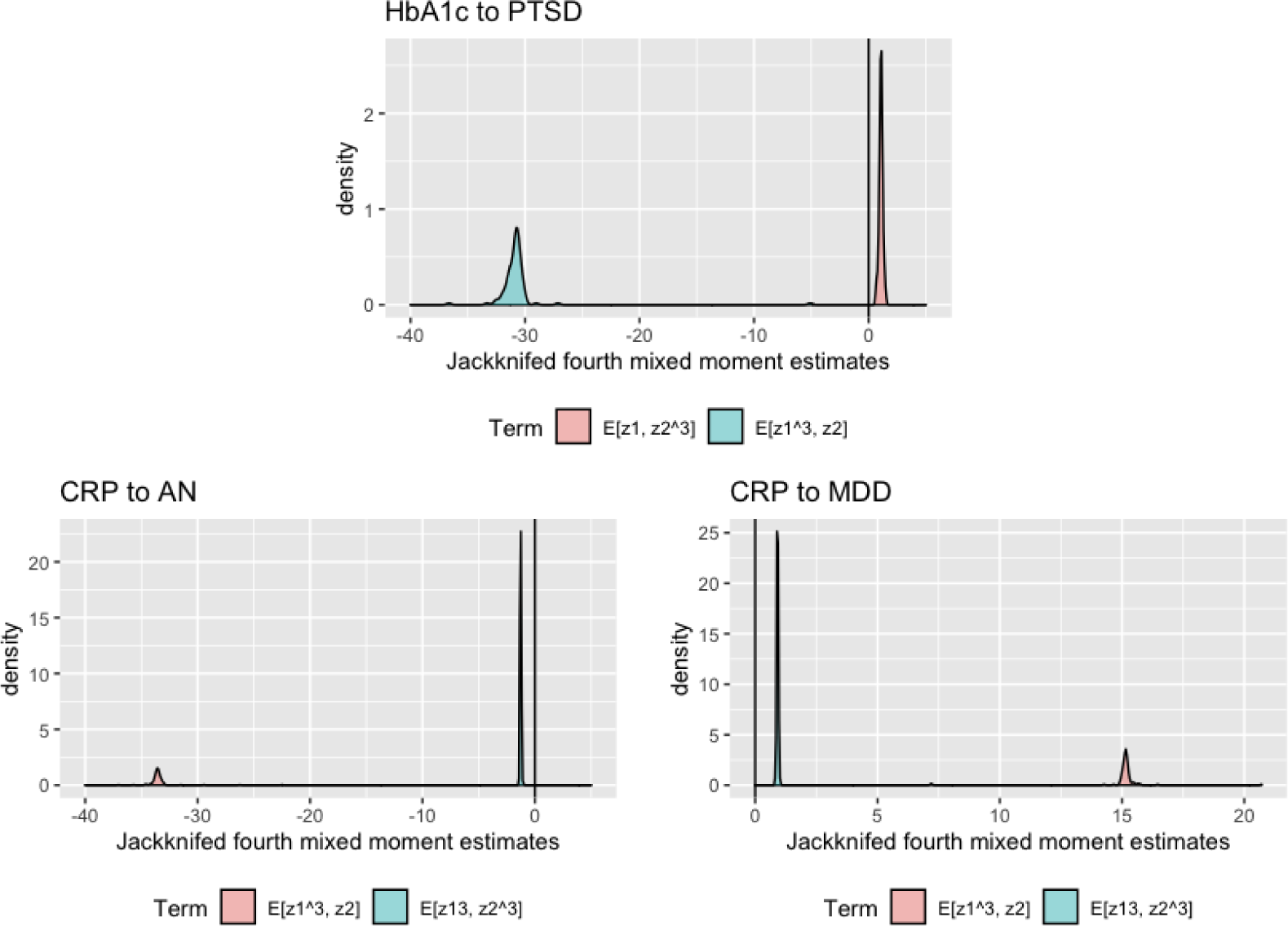
Analysis of an LCV model violation in the HbA1c to PTSD model. As outlined in the supplementary text, we demonstrated that the two mixed fourth moments (cokurtosis) estimates for HbA1c to PTSD were opposite in sign, with an outlier jackknifed estimate of which suggests discordant SNP effects relative to the genome-wide signal. Conversely, the CRP to AN and CRP to MDD had consistent signs of and relative to each other and the genetic correlation. In both CRP models, which implies GCP > 0.

**Supplementary Figure 2.**
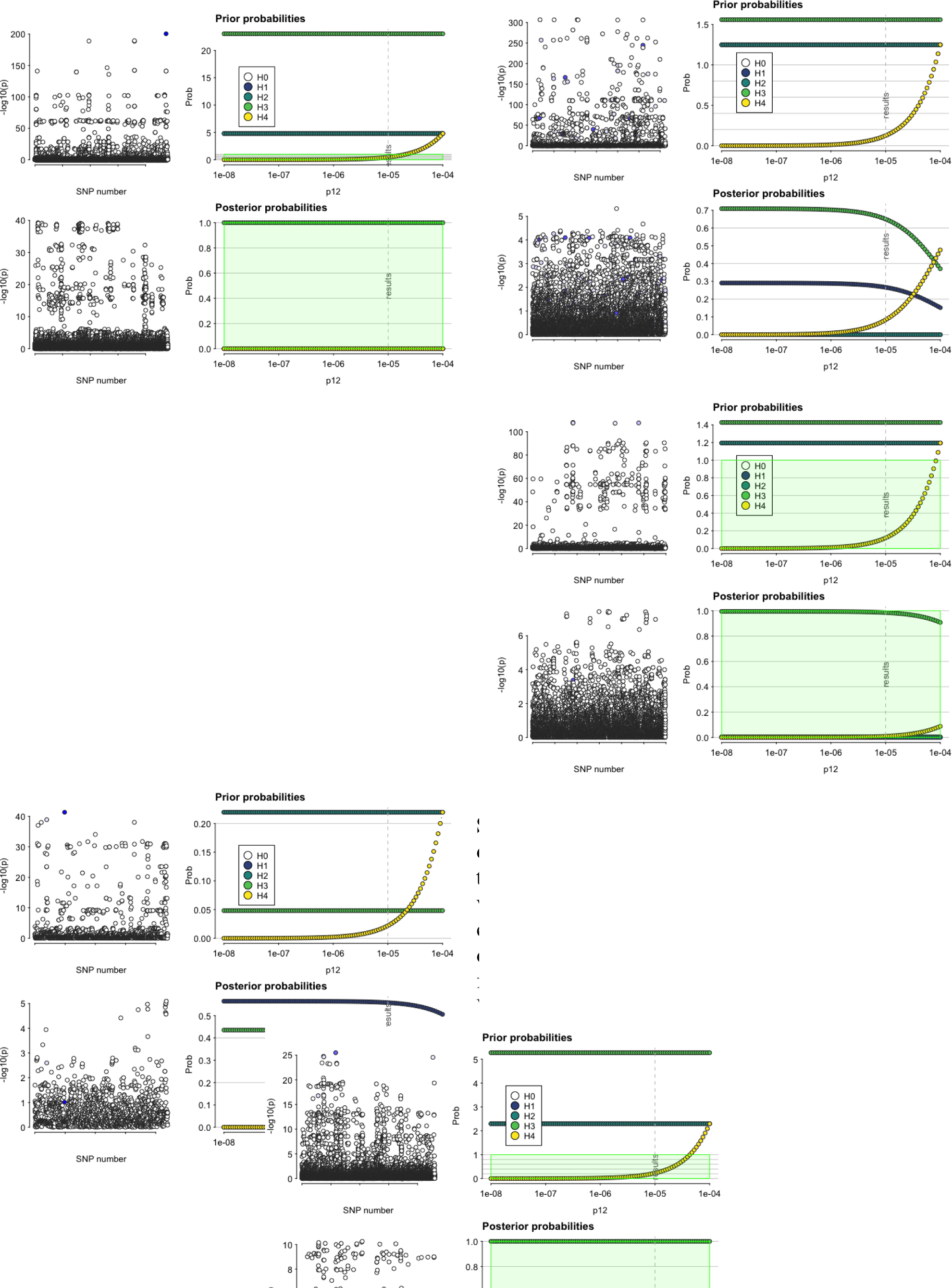
The effect of different prior probabilities for the hypothesis that the SZ and CRP share a same causal variant. Each of these plots represent the colocalisation results for the five regions outlined in supplementary table 24, in order from region 1 to 5, left to right, top to bottom. We plot the effects of varying the default prior probability for the shared causal variant above or below its default of 1 x 10^-5^.

**Supplementary Figure 3.**
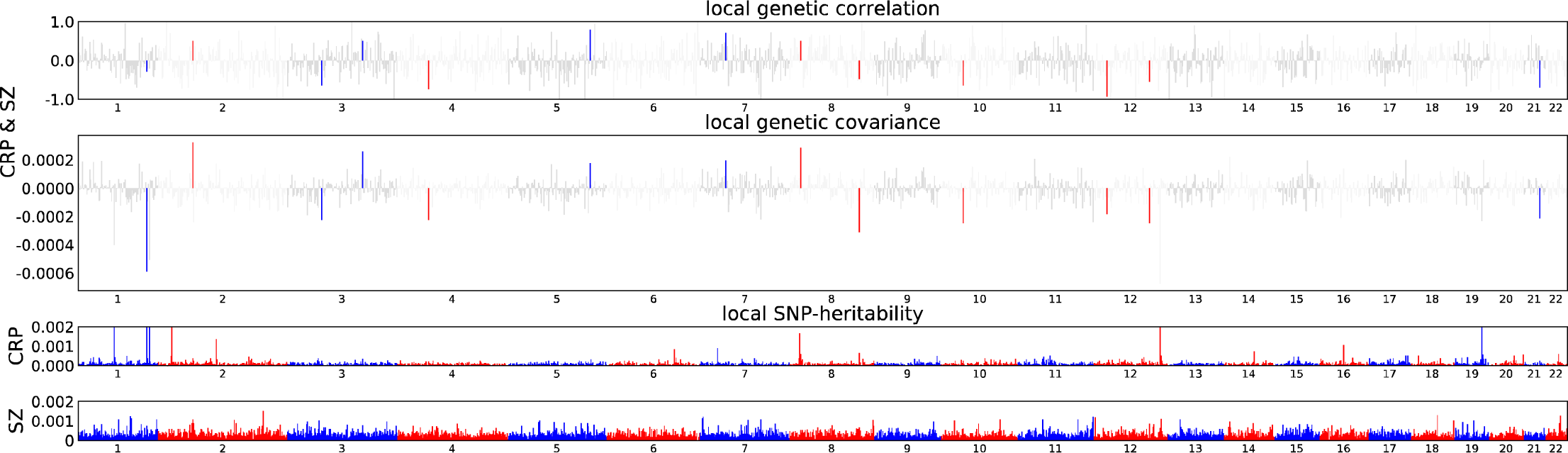
Local genetic covariance and correlation between schizophrenia and CRP as estimated by ρ-HESS. The first two panels are estimates of local genetic correlation and covariance for each approximately independent LD block throughout the genome. Coloured bars represent significant estimates. The remaining two panels are local estimates SNP heritability in those same regions for CRP and SZ, respectively.

## Notes

### Competing Interest Statement

The authors have declared no competing interest.

### Author Declarations

University of Newcastle Human Research Ethics Committee (HREC)

