## Supplementary Text for "Genetic estimates of correlation and causality between blood-based biomarkers and psychiatric disorders"

**Contents**

Psychiatric genome-wide association studies	2

Blood-based biomarker GWAS in the UK biobank (Neale group)	5

Description of the Australian Schizophrenia Research Bank (ASRB) cohort	5

Mendelian randomisation methodology	7

Model violations in the latent causal variable (LCV) approach	15

Colocalisation analyses between schizophrenia and C-reactive protein	16

Supplementary Text References	17

**Psychiatric genome-wide association studies**

We outline some basic information regarding each of the psychiatric GWAS included in this study, with more extensive descriptions available in the referenced original studies:

*ADHD*

The ADHD GWAS was a meta-analysis of primarily European cohorts, however, to accurately estimate genetic correlation by LDSR, in this study we only utilised the European subset of the meta-analysis. As a result, the GWAS was performed on 19,099 ADHD cases and 34,194 controls. The SNP heritability for ADHD on the liability scale was 21.6% assuming a population prevalence of 5%, with 12 genome-wide significant loci discovered in the overall meta-analysis. Further details can be found in the publication describing the GWAS (1).

*Anorexia nervosa*

Anorexia nervosa was subjected to a GWAS using a meta-analysis of European ancestry cohorts that totalled 16,992 cases of anorexia nervosa and 55,525 controls, with eight genome-wide significant loci uncovered. Assuming a conservative population prevalence of 0.9%, the SNP heritability was around 11%. Further details can be found in the publication describing the GWAS (2).

*Autism spectrum disorder*

The autism GWAS was a European ancestry meta-analysis composed of 8,381 cases and 27,969 controls that identified five genome-wide significant loci. SNP heritability for ASD was approximately 11.8% on the liability scale, assuming a 1.2% prevalence. Further details can be found in the publication describing the GWAS (3).

*Bipolar disorder*

The bipolar disorder GWAS included 20,352 cases and 31,358 controls, with 19 genome-wide significant variants uncovered in the discovery GWAS. SNP heritability for BIP was estimated as ~ 17% given a 0.5% population prevalence. Further details can be found in the publication describing the GWAS (4).

*General cognitive ability*

General cognitive ability (cognition) was subjected to GWAS by meta-analysis of cohorts totalling 269,867 individuals, the phenotypes were a range of neurocognitive tests assessing variables primarily related to fluid intelligence, with a general *g* factor of intelligence extracted for the majority of cohorts as described in detail in the supplementary material of the GWAS manuscript (5). SNP heritability in the full sample was estimated as 19%, with 242 genome-wide significant loci.

*Major depressive disorder*

The major depressive disorder GWAS was a European ancestry meta-analysis, however, given 23andMe contributed data to this study, the entire GWAS cohort was not publicly released as summary statistics, with full details given in the Wray *et al.* publication (6). Excluding the 23andMe cohort, 59,851 cases and 113,154 controls were available for analysis, with SNP heritability for this subset of the cohort reported as approximately 10% (15% population prevalence), with five genome-wide significant loci identified from this subset without the 23andMe cohort.

*Obsessive compulsive disorder*

The OCD GWAS was comprised of 2688 cases and 7037 controls. There were no genome-wide significant loci identified in this study, with full details in the original publication (7).

*Post-traumatic stress disorder*

The PTSD GWAS was a trans-ancestral meta-analysis, however, as with ADHD we only utilised the European subset due to methodological constraints in estimating genetic correlation from trans-ancestral samples with LDSR. There were 23,212 European ancestry cases, 151,447 controls. SNP heritability was estimated at around 4% given a 10% prevalence, with two genome-wide significant loci discovered in the European subset. Full details are provided in the PTSD GWAS publication (8).

*Schizophrenia*

Analogous to ADHD, the PGC3 schizophrenia GWAS was majority European ancestry, however, we utilised the European subset as with the other trans-ancestral GWAS (total European sample size = 130,644), with a SNP heritability estimate of 24%. Full details are outlined in the preprint of the PGC3 GWAS (9).

*Tourette’s syndrome*

The Tourette syndrome GWAS was performed on 4819 cases and 9488 controls of European ancestry. A single genome-wide significant locus was uncovered, with SNP heritability estimated as 21% given a 0.8% prevalence, further details are outlined in the GWAS publication (10).

**Blood-based biomarker GWAS in the UK biobank (Neale group)**

The biochemical GWAS interrogated to estimate genetic correlation and infer potential causal relationships were a collection of GWAS of blood based biochemical traits collected from UK biobank participants, including, lipids, hormones, blood cell counts, and vitamins. These GWAS were performed by Ben Neale’s group and were publicly available for download from the group’s website (<http://www.nealelab.is/uk-biobank>). Specifically, we selected 50 of these traits for which the estimate of SNP heritability was significantly different from zero and categorised as ‘high’ or ‘medium’ confidence estimates. The full details of these categorisations have been outlined elsewhere (<https://nealelab.github.io/UKBB_ldsc/confidence.html>). In all instances, we utilised the GWAS summary statistics for which the phenotype of interest was subject to inverse-rank normal transformation. The covariates for these GWAS were age, age^2^, sex, age x sex, age^2^ x sex, and the first 20 principal components.

**Description of the Australian Schizophrenia Research Bank (ASRB) cohort**

Full details regarding the ASRB cohort have been described previously, we briefly outline the analyses relevant to this study below. The ASRB is public bank of clinical and cognitive data, including structural magnetic resonance imaging (MRI) scans and DNA samples, for a large cohort of schizophrenia cases and controls, collected across five cooperating sites in Australia, as outlined elsewhere (11).

*Genotyping*

Investigators were blinded to phenotype during processing of samples for genotyping and/or sequencing. Genomic DNA from peripheral blood mononucleocytes was extracted for SNP genotyping and/or WGS. SNP genotyping utilised the Illumina Infinitium Human 610K (610-Quad) BeadChip in accordance with standard manufacturer protocols. The quality control procedure for samples genotyped by SNP array in the ASRB cohort are as follows. Before imputation genotyped autosomal SNPs were removed if they failed a series of quality control (QC) criteria implemented with PLINK 1.9: MAF < 0.01, call rate < 98% and Hardy-Weinberg Equilibrium (HWE) *P* < 10^-6^. Ambiguous SNPs not assigned to a strand were also filtered out along with all non-autosomal sites. Individuals with discordant sex information were excluded from the cohort along with one individual of a pair with an Inbreeding Coefficient (IF) > 0.2. High quality autosomal SNPs (MAF > 0.05) in relative linkage equilibrium (R^2^ < 0.1) were input for relatedness testing, whilst regions of long-range linkage disequilibrium were removed. Individuals with high relatedness defined by genome wide identity by state ($\hat{\pi}$ > 0.185) were also removed. Principal components analysis (PCA) was also undertaken with PLINK 1.9. Population outliers were excluded using k-means clustering, wherein the five generated clusters represented the super-populations in the 1000 Genomes. After phasing haplotypes with 10 megabase chunks and 500 kilobase (kb) overlap with Eagle 2.3.2, imputation was performed on loci which passed QC using Minimac3 default parameters and the 1000 Genomes Phase 3 European reference panel. High-quality SNPs post-imputation were retained for analysis (INFO score > 0.8, missingness < 2%, N_SNPs_ = 7,199,582).

*Cognitive subtyping*

As described in Green *et al.,* the multidimensional Grade of Membership (GoM) clustering technique was used to derive subgroups of cognitive performance in the ASRB schizophrenia cohort with available cognitive data (12). Briefly, nine cognitive measures were utilised as input into the GoM – Wechsler Test of Adult Reading, Wechsler Abbreviated Scale for Intelligence, Controlled Oral Word Association Test, Letter Number Sequencing (Scaled Score), Repeatable Battery for the Assessment of Neuropsychological Status – immediate memory, Repeatable Battery for the Assessment of Neuropsychological Status – constructional, Repeatable Battery for the Assessment of Neuropsychological Status – language, Repeatable Battery for the Assessment of Neuropsychological Status – attention, Repeatable Battery for the Assessment of Neuropsychological Status – delayed memory, and Repeatable Battery for the Assessment of Neuropsychological Status – total scaled score. The most parsimonious GoM model partitioned schizophrenia cases into the CD subtype, with a greater degree of global cognitive impairment, and a cognitively spared (CS) subtype whose performance was intermediate to the CD subtype and healthy controls. In the genotyped cohort this resulted in 170 individuals with schizophrenia subtyped as CD, and 221 subtyped as CS. The CD cohort was majority male – CD: 63% male, whilst the CS cohort was majority female (65%).

*Polygenic score generation*

Autosomal polygenic scores were generated using the default settings of PRSice v 2.3.3 linux with a *P* value threshold of *P* < 0.05. These were scaled in the subsequent CD vs CS cohort to have mean zero and unit variance.

**Mendelian randomisation methodology**

Given that the MDD and AN GWAS had some sample overlap with the UK biobank, we utilised a different CRP GWAS to select instrumental variables in the Mendelian randomisation analyses (MR). Specifically, this GWAS was a meta-analysis of up to 204,402 European ancestry individuals imputed to the HapMap3 panel, with each study adjusting for age, sex, and population structure, as described in Lighthart *et al.* (13). We note that ‘ratio estimates’ described forthwith relate to the effect of an IV-SNP on the outcome divided by the effect of an IV-SNP on the exposure variable.

*Univariable Mendelian randomisation*

The primary IV assumptions in MR are as follows: variant must be rigorously associated with the exposure; the variant must be independent of all confounders of the exposure-outcome relationship, and the variant must be associated with the outcome only by acting through the exposure. The methods we employed in this study make varying assumptions regarding the validity of IVs and are supplemented with sensitivity analyses as described below. Moreover, we outline the results for each of the CRP $\to$ psychiatric univariable models.

Strength of the CRP IVs

Using the default stringent pruning criterion such that genome-wide significant CRP SNPs were in linkage equilibrium, and thus, independent (*r*^2^ < 0.001), there were 50 IVs available in the OCD, MDD, and SZ GWAS, whilst 49 were available in the AN GWAS. During the harmonisation step, we removed all potential palindromic IV SNPs. The *F* statistic for either the 49 or 50 IVs was not materially different (49 IVs = 151.3068, 50 IVs = 149.1304), whilst the *I*^2^ statistic was large enough such that MR-Egger could be performed (*I*^2^ > 0.9).

Inverse-variance weighted (IVW) estimator

The IVW estimator is our primary model and is considered to be the most well-powered MR method (14). Briefly, the pooled causal effect of the *j* IV exposure effects (*X*) on the outcome (*Y*) are estimated, where σ is the IV-outcome standard error (equation 1).

$$\hat{\beta}_{IVW}= \frac{\sum_{j} \hat{\beta}_{Yj}\hat{\beta}_{Xj} \sigma_{Yj}^{-2}}{\sum_{j} \hat{\beta}_{Xj}^{2} \sigma_{Yj}^{-2}} [1]$$

We used the IVW estimator with multiplicative random effects, rather than fixed effects, as this for multiplicative overdispersion in the causal estimates, and is likely more precise in the presence of heterogeneity that could be indicative of confounding pleiotropy. We note that the MR estimate using the fixed or random effects approach will be identical, but the standard errors will differ. The key limitation of the IVW approach is that it has a zero percent breakdown point, in other words, a single invalid IV may bias the overall estimate – necessitating the use of other MR methods to evaluate the rigour of the causal estimate to different underlying assumptions regarding IV validity. One method to address is this is to utilise robust regression for the IVW estimate to provide additional guarding against outliers and penalisation of the heterogenous weight estimators using Cochran’s *Q* statistic, results for which are reported in supplementary table 21 (15).

In the univariable CRP analyses, we found that that there was statistically significant evidence of a protective effect of CRP on SZ and AN, whilst the MDD and OCD estimates were not significantly different than zero. As expected, the fixed effects IV estimate yielded lower standard errors and more significant *P* values, however, we assert the estimator with multiplicative random effects is more suitable given the number of IVs and complexity of the phenotypes in question. Using robust IVW with or without weights penalised for heterogeneity did not materially alter the results.

Median based estimators (majority valid assumptions).

An alternative to the IVW approach are median based estimators which do not assume that all IVs will be valid (16). This arises because median based estimators take the median, rather mean as in IVW, of ratio estimates. We utilise two median based estimators in this study which are subject to the majority valid assumption – the simple and weighted median estimators. Specifically, the simple median denotes the median ratio estimate and is valid so long as less than half of the IVs are invalid, however, this method is inefficient in the presence of ratio estimates that vary considerably. As a result, the weighted median estimator, whereby more precise estimates are upweighted, is said to be unbiased so long as < 50% of the weight comes from invalid IVs. Analogous to the IVW models, we also included a penalised weighted median estimator, whereby the contribution of heterogenous ratio estimates is down-weighted.

The CRP $\to$ SZ estimate remained statistically significant using a weighted and penalised weighted median estimator, however, the estimate only trended towards statistical significance (*P* = 0.07) using the less powerful simple median estimator. Moreover, the CRP → AN estimate across the three median estimators remained relatively consistent, however, were not statistically significant.

Plurality valid (modal estimators and the contamination mixture model)

A complementary approach to the median based estimators are MR methods which make the related ‘plurality valid’ assumption – which assumes out of groups of IVs having the same asymptotic estimate, the largest group will be comprised of valid IVs. We implement two approaches will make this assumption – a weighted mode estimator and contamination mixture models (17,18). The mode-based approach does not take the mode of the direct ratio estimates itself, as these ratio estimates will not be identical, rather the mode of the smoothed empirical density function of each ratio estimate. The contamination mixture model also implies the plurality valid assumption Specifically, a likelihood function based on IV effects is specified from a product of two component mixture distributions (IV is valid and IV is invalid with distributional assumptions described by Burgess *et al.* (18)). Contamination mixture models are useful as it can identify groups of similar ratio estimates which may be indicative of multiple causal mechanisms. The causal estimate represents that exposure-outcome estimate that maximises the profile likelihood, whilst the confidence interval can have multiple disjoint ranges. To implement the contamination mixture model, we utilised a variety of ψ values, with ψ as a multiple of the ratio estimate standard error, to define the standard deviation of invalid IVs, higher values of ψ means that the assumed standard deviation of invalid IV estimates is larger. We utilised ψ = 0.8, 1, 1.2, 1.5, and 1.8 in this study as recommended in the contamination mixture model manuscript.

The CRP $\to$ SZ estimate remained statistically significant using the weighted mode estimator with each 1 ln mg/L increase in CRP associated with a ~ 22% reduction in the odds of SZ: OR = 0.88 [95%: 0.83, 0.94], *P* = 1 x 10^-4^. Similarly, the causal estimate was significant in the contamination mixture models irrespective of the ψ parameter utilised. CRP $\to$ AN was directionally consistent (negative) but only trended towards significance using the weighted mode estimator (*P* = 0.10). There was evidence of significant causal effect when ψ < 1.2 in the contamination mixture models, however, the estimate only trended towards significance when the standard deviation of invalid IVs was assumed to be higher (ψ $\geq$ 1.2).

MR-Egger (InSIDE assumption)

An MR egger model was then constructed; an adaption of Egger regression wherein the exposure effect is regressed against the outcome with an intercept term ($\theta_{0}$) added to represent the average pleiotropic effect (equation two) (19).

$$\hat{\beta}_{Yj} \sim\theta_{0}+ \theta_{1}\hat{\beta}_{Xj}+ \varepsilon_{j} [2]$$

The key assumption of the MR egger model is referred to as Instrument Strength Independent of Direct Effect (InSIDE), which assumes that there is no significant correlation between direct IV effects on the outcome and genetic association of IVs with the exposure. In other words, the InSIDE assumption is violated if pleiotropic effects act through a confounder of the exposure-outcome association(19–21). We also tested whether the Egger intercept is significantly different from zero as a measure of unbalanced pleiotropy or violation of the InSIDE assumption. The robust regression methodology and Cochran’s *Q* based penalisation of ratio estimate weights were also applied the MR-Egger estimates.

Once more, CRP demonstrated a consistent and statistically significant protective effect on SZ using the MR-Egger model with or without the robust or penalisation alterations. Furthermore, the intercept from any of these three models was not significantly different from zero, and therefore, does not provide evidence that there is an effect of unbalanced pleiotropy. The CRP to AN estimate using Egger methods were all negative but not statistically significant. The Egger intercepts were also not significantly different from zero, however, there was a trend towards this (*P* = 0.07), and thus, there could be some nominal evidence of unbalanced pleiotropy.

Sensitivity analyses of heterogeneity and pleiotropy

In addition to the Egger intercept tests described above, we also evaluated statistical evidence of confounding pleiotropy by assessing the heterogeneity of ratio estimates (Cochran’s *Q* and MR-PRESSO), as well as a related test which iteratively removes one IV at a time and recalculates the IVW estimate (leave-one-out-analyses) (22,23). MR-PRESSO is underpinned by the residual sum of squares (RSS), which serves as a heterogeneity measure of ratio estimates. Specifically, an IVW estimate using the IVs is calculated in a leave-one out fashion; if the RSS is decreased significantly relative to a simulated Gaussian distribution of expected RSS, then that variant is excluded from the IVW model. The MR-PRESSO global test of pleiotropy compares the summed observed vs expected RSS, that is, more heterogeneity than expected by chance. Furthermore, we also tested heterogeneity between the IV effects using Cochran’s *Q* statistic.

The MR-PRESSO and Cochran’s *Q* estimates were indicative of statistically significant heterogeneity amongst the ratio estimates for the AN and SZ models, which may signal confounding pleiotropy, or could be a result of biological heterogeneity of the effect of CRP on these outcomes. Crucially, the MR PRESSO outlier corrected estimate, whereby heterogeneous ratio estimates were removed, remained significant for both AN and SZ. Similarly, removing each IV iteratively did not greatly affect the CRP estimate on AN or SZ, regardless of which IV was removed. As outlined in supplementary table 19, there were four instances whereby the removal of an IV rendered the model nominally not significantly different from zero, however, *P* < 0.08 for all instances upon the removal of said IV. The outlier in the SZ model was an upstream SNP of CRP, and thus, is plausible that this represents a biologically relevant direct effect on CRP. In the AN model, two of the outliers seem plausibly directly related to CRP, however, one outlier in the *HNF1A* gene was also associated with many other biochemical traits, and thus, this could be an instance of confounding pleiotropy and warrants further investigation.

*Multivariable Mendelian randomisation*

Multivariable Mendelian randomisation is an extension of the univariable MR approach which allows the estimate of the direct effect of an exposure on an outcome conditioned on other exposures. We constructed multivariable models to examine whether the univariable total estimates of CRP on the psychiatric outcomes were confounded or mediated by two closely related variable to CRP biology – body mass index (BMI) and interleukin-6 signalling (IL-6 and IL-6R). The BMI and IL-6 GWAS chosen were both non-UKBB samples, as with the CRP GWAS. Specifically, the BMI GWAS was a meta-analysis of up to 322,154 European individuals, with BMI residualised through regression on age, sex, and principal components for the GWAS phenotype, although various differing methodologies were applied in each cohort which contributed to the meta-analysis, as described in the GWAS manuscript (24). We also used measured protein expression of IL-6 and its receptor IL-6R from a plasma study using 3,394 participants from the IMPROVE study, that is, individuals with risk factors for coronary artery disease but without symptoms (25). Log_10_-transformed protein levels were regressed on age, sex, recruitment centre, protein analysis batch, smoking, diabetes, and hypertension at baseline – with the subsequent standardised residuals subjected to GWAS, as outlined in original study (26).

We sought to assess the strength of candidate IVs in the following three multivariable models – CRP, IL-6 and BMI, CRP and BMI, and CRP, IL-6R, and BMI using the conditional *F* statistic described by Sanderson *et al.*, which tests whether the IVs strongly predict each exposure, conditional on the other exposures in the model (27,28). Firstly, we attempted to select genome-wide significant SNPs for each exposure such that the conditional *F* statistic > 10 – whereby SNPs which are genome-wide significant for at least one exposure are included in the model. The required strength for the *F* statistic was satisfied, except in the CRP/IL-6/IL-6R model, whereby IL-6 and IL-6R were weak. To address this, we relaxed the inclusion criterion in this model only to suggestive associated SNPs (*P* < 1 x 10^-5^) – the conditional *F* statistic for IL-6R then exceeded 10 (Supplementary Table 23), however, IL-6 was still marginally below 10 (*F* = 9.93). Given that weak instruments in a two-sample setting are likely to bias the estimate towards the null (27), we proceeded to estimate the direct effect of IL-6 even though it is underpowered. Future larger studies of IL-6 with genome-wide summary statistics available are needed to address this.

Four MVMR models were implemented in this study – a multivariable IVW estimator with multiplicative random effects (mvIVW), a LASSO-type penalisation approach (mvLASSO), a median based estimator (mvMedian), and multivariable extension of MR-Egger (29). The assumptions regarding the IVW, median, and Egger models have already been extensively outlined in this supplement. The regularisation mvLASSO approach, as outlined by Grant and Burgess (29), adds an intercept term ($\theta_{0}$) for each IV-variant and applies LASSO type penalisation such that the intercepts are shrunk towards zero for valid IVs. The tuning parameter (λ) controls how many variants are deemed invalid, and thus, shrinkage is not applied to their intercept term – with a data driven approach utilised to select λ. The corresponding estimates from each of these multivariable models can be defined as the effect of a unit increase of the exposure on the outcome, conditioned on the remaining exposures.

**Model violations in the latent causal variable (LCV) approach**

As described in the results of the manuscript, in the LCV model constructed for HbA1c $\to$ PTSD, we observed an unusual phenomenon whereby the posterior mean GCP strongly indicated partial genetic causality of HbA1c on PTSD ($\hat{GCP}$ = 0.76), whilst its *Z* score was negative (*Z* = -6.51), which implies the opposite direction of partial genetic causality from PTSD to HbA1c. To reconcile this, we investigated the raw estimates of the mixed fourth moments (cokurtosis) for HbA1c (*Z*_1_) and PTSD (*Z*_2_) and the genetic correlation which was strongly significantly greater than zero ($r_{g}$ = 0.21). The full methodology of the LCV model is described in detail by O’Connor and Price, however, we focus here on the cokurtosis estimates (30). We found that that $E\left( Z_{1}^{3} Z_{2} \right)$was strongly negative, with a peak in the distribution each of the 100 jackknifed block estimates at around -30, whilst $E\left( Z_{1}Z_{2}^{3} \right)$ was positive in the majority of the jackknifed cokurtosis estimates, with one block that was marginally negative (-4), suggesting the existence of SNPs with discordant effects than the genome-wide signal. Given that $\left| E\left( Z_{1}Z_{2}^{3} \right) \right| \geq\left| E\left( Z_{1}Z_{2}^{3} \right) \right|$implies that $\hat{GCP}$ > 0, as was the case here, it is indicative of an effect from HbA1c to PTSD. However, due to the discordant signs of the cokurtosis estimates relative to each other and the genetic correlation, as well as evidence of an outlier jackknifed $\left| E\left( Z_{1}Z_{2}^{3} \right) \right|$ less than zero the model is unstable. We represent this graphically in supplementary figure 1 – whereby the jackknifed cokurtosis estimates for the unstable HbA1c $\to$ PTSD model, are contrasted to CRP $\to$ AN, and CRP $\to$ MDD, which behave as expected. HbA1c is a complex phenotype that is influenced by both haematological and glycaemic factors, and thus, it is plausible that loci affecting different aetiological components of this phenotype, may have discordant effects relative to the genome-wide signal. As a result, further investigation of the relationship between HbA1c and PTSD is required to resolve whether the genetic correlation reflects horizonal pleiotropy and/or a causal relationship.

**Colocalisation analyses between schizophrenia and C-reactive protein**

We sought to identify specific instances of genetic overlap between the UKBB CRP and PGC3 schizophrenia GWAS. Specifically, colocalisation analyses were applied for lead SNPs identified by the PGC in the full trans-ancestral SZ GWAS which were also genome-wide significant in the CRP GWAS. The *coloc* package for colocalisation is a Bayesian approach which the configuration of approximated Bayes’ factors assuming a single causal variant for each SNP in the region is assessed to calculate the posterior probability of the following hypotheses, as described in Giambartolomei *et al.*(31):

H_0_: no association at this locus with either trait,

H_1_: association only with trait 1,

H_2_: association only with trait 2,

H_3_: locus associated with both traits, however, different underlying causal variant, and

H_4_: locus associated with both traits, both sharing the same causal variant.

We subjected each locus for which colocalisation analyses were performed to a sensitivity analyses where we calculated the posterior probability (*PP*) of H_4_ using varying priors for that hypothesis (Supplementary Figure 2).

**Supplementary Text References**
